# SARS-CoV-2 infection in Health Care Workers in a large public hospital in Madrid, Spain, during March 2020

**DOI:** 10.1101/2020.04.07.20055723

**Authors:** María Dolores Folgueira, Carmen Muñoz-Ruipérez, Miguel Ángel Alonso-López, Rafael Delgado, on behalf of the Hospital 12 de Octubre COVID-19 Study Groups

## Abstract

**Background:** On January 31^st^ the first case of COVID-19 was detected in Spain, an imported case from Germany in Canary Islands, and thereafter on February 25^th^ the first case was detected in Madrid. The first case of COVID-19 was confirmed at the Hospital Universitario 12 de Octubre on March 1^st^, a large public hospital with 1200 beds, covering an area over 400000 inhabitants in southern Madrid. During March 2020 highly active circulation of SARS-CoV-2 was experienced in Madrid with 24090 cases officially reported by March 29^th^.

**Methods:** Since the beginning of the epidemics the Occupational Health and Safety Service (OHSS) organized the consulting and testing of the hospital personnel with confirmed exposure and also those presenting symptoms suggestive of viral respiratory infection. For molecular diagnosis of SARS-CoV-2 infection both nasopharyngeal and oropharyngeal swabs were obtained from suspected cases and processed at the Microbiology Laboratory by automatized specific PCR methods that was operative from February 25^th^ as part of the preparedness.

**Results:** From a total of 6800 employees of the hospital, 2085 (30,6 %) were tested during the period 1-29 March 2020, some of them repeatedly (2286 total samples). The first HCW infected was confirmed on March 9^th^. A total of 791 HCW and personnel were confirmed to be infected by March 29^th^, representing 38% of those tested and 11,6 % of all the hospital workers. The proportion of infected individuals was estimated among the different groups of occupational exposure and the evolution of the cases during the expansive epidemic wave was compared between HCW and those patients attending at the Emergency Department (ER) during the same period and adjusted by the same age range. There were no statistically significant differences in the proportion of SARS-CoV-2 positive PCR detection between HCW from high risk areas involved in close contact with COVID-19 patients in comparison with clerical, administrative or laboratory personnel without direct contact with patients. The curves of evolution of accumulated cases between patients and HCW during March 2020 showed an almost parallel shape.

**Discussion:** The recommendation from our OHSS did not include testing of asymptomatic cases but was highly proactive in testing even patients with minor symptoms therefore, a high proportion of HCW and non-sanitary personnel was tested in March 2020 during the rapid period of expansion of the epidemics in Madrid, accounting for a total of 30,6 % of the hospital employees. Most of the COVID-19 cases among the hospital HCW and personnel were mild and managed at home under self-isolation measures, however 23 (3%) required hospitalization mostly due to severe bilateral interstitial pneumonia, two of those cases required mechanical ventilation at the ICU. No fatalities occurred during the study period.

Although there were some cases of highly probable transmission from COVID-19 patients to HCWs, mainly at the first phase of the epidemics, there were no significant differences on the infection rates of HCW and hospital personnel that can be related to working in areas of high exposure risk. Furthermore, the evolution of cases during the same time period (March 2020) between patients attending the ER and hospital staff suggests that both groups were driven by the same dynamics. This experience is similar to the communicated from Wuhan verified by the WHO Joint Mission and also from recent experiences at hospital in the Netherlands, where most of the infections of HCW were related to household or community contacts.

**Significance:** Since the collective of hospital HCW are exhaustively screened in specific centers, their rate of infection for SARS-CoV-2 could be an indicator of the epidemic dynamics in the community. There appears to be a close connection between HCW infection and the driving forces of transmission in the community. Although we cannot exclude an additional risk factor of infection by SARS-CoV-2 due to the fact of the hospital environment, the similar proportions of positive cases among all the areas of the hospital and the evolutive wave of infection, as compared with the community, are clear arguments against a major factor of occupational risk. Exhaustive testing, such as the one carried out in our institution, covering over one third of all the workers, could be used as a reference of the population infected in the community. Since a significant proportion of COVID-19 cases can be asymptomatic and not all the hospital employees were actually tested, it is highly likely that this 11,6 % is a minimum estimation of the impact of SARS-CoV-2 circulation in Madrid during the first 4 weeks of the epidemics. This is in high and clear contrast with the official figures circulating at national and international levels. This has important implications to more precisely estimate the actual number of cases in the community and to develop public health policies for containment, treatment and recovery.

## Introduction

COVID-19 is an acute respiratory tract infection caused by a new human coronavirus SARS-CoV-2(1) that emerged in Wuhan, China, in late 2019. On January 31^st^ the first case of COVID-19 was detected in Spain, an imported case from Germany in Canary Islands, and thereafter on February 25^th^ the first case was detected in Madrid(2). The first case of COVID-19 was confirmed at the Hospital Universitario 12 de Octubre on March 1^st^, a large public hospital with 1200 beds, covering an area over 400000 inhabitants in southern Madrid. The hospital has currently 6800 employees in all professional areas both health care workers (HCW) along with laboratory, clerical and administrative personnel. During March 2020 highly active circulation of SARS-CoV-2 has been experienced in Madrid with 24090 cases officially reported by March 29^th^.

## Methods

Since the beginning of the epidemics the Occupational Health and Safety Service (OHSS) organized the consulting and testing of the hospital personnel with confirmed exposure and also those presenting symptoms suggestive of viral respiratory infection. For molecular diagnosis of SARS-CoV-2 infection both nasopharyngeal and oropharyngeal swabs were obtained from suspected cases and processed at the Microbiology Laboratory by automatized extraction and specific PCR methods that was operative from February 25^th^ as part of the preparedness for the potential epidemic spread. Samples were collected with flocked swabs in UTM™ viral transport medium (Copan Diagnostics, Brescia, Italy). After an external lysis, nucleic acid extraction was performed in the MicrolabStarlet IVD platform using the STARMag 96 × 4 Universal Cartridge Kit (Seegene, Seoul, South Korea) or in the NucliSENS EasyMAG instrument (bioMerieux, Marcy l’Etoile, France). For rRT-PCR, we used the LightCycler 480 System instrument II (Roche Life Science, Indianapolis, IN, USA), performing the test TaqMan 2019nCoV assay Kit v1, provided by Thermosfisher Scientific, that amplifies three different viral regions in singleplex reactions(3). Appropriate controls (positive and negative) were tested in each run, as well as an internal control to rule out the presence of PCR inhibitors in the samples. For comparison of different occupational risk, we established three different groups of HCW. High risk exposure when the worker belonged to the emergency room or hospital areas where they have concentrated patients with COVID19, as well as areas of intensive care and resuscitation. Medium risk exposure was defined for those workers who were in occasional contact with patients with COVID19, such as surgery, Oncology, Hematology, Radiology, Ob/Gyn, Pediatrics and Medical areas non- COVID19 related. Low risk exposure included workers who were not in contact with patients, such as laboratory, Pharmacy, Kitchen and administrative personnel. Graph Pad Prism V7 was used for statistical analysis.

Ethical approval: This study has been approved by the Institutional Internal Review Board (IRB) of Hospital Universitario 12 de Octubre: N-CEIm: 20/169, April 5^th^ 2020.

## Results

From a total of 6800 employees of the hospital, 2085 (30,6 %) were tested during the period 1-29 March 2020, some of them repeatedly (2286 total samples). The first HCW infected was confirmed on March 9^th^. A total of 791 HCW were confirmed to be infected by March 29^th^, representing a 38% of those tested and an 11,6 % of all the hospital workers. Most of the COVID-19 cases among HCW were mild, however 21 infected HCW (2,6 %) required hospitalization mostly due to moderate to severe bilateral interstitial pneumonia. Two of those cases required mechanical ventilation at the ICU. No fatalities occurred during the study period.

The proportion of infected individuals was estimated by the different groups of occupational exposure and the evolution of the cases during the expansive epidemic wave was compared between HCW and those patients attending at the ER Department during the same period and adjusted by the same age range of HCW (20-68 years old) (Figure 1). HCW were classified according to their exposure to infected patients and/or aerosol generation. Three levels of risk were defined for most of the HCW studied: There were no statistically significant differences in the proportion of SARS-CoV-2 positive PCR detection between HCW from high risk areas involved in close contact with COVID-19 patients in comparison with intermediate or low risk areas (Table 1). The curves of evolution of accumulated cases between patients and HCW during March 2020 showed an almost parallel shape (Figure 1).

**Table 1:** HCW were classified according to their exposure to infected patients and/or aerosol generation. Three levels of risk were defined for the different areas shown in the table. Nor all the areas are shown (601 out of 791 + HCW and Employees): High risk exposure when the worker belonged to the emergency room or hospital areas where they have concentrated patients with COVID19, as well as areas of intensive care and resuscitation. Medium risk exposure was defined for those workers who were in occasional contact with patients with COVID19, such as surgery, pediatrics and medical areas non-COVID19 related. Low risk exposure included workers who were not in contact with patients, such as laboratory and administrative personnel. Proportion of positive PCR HCW is shown as %. A two tailed Fisher test (Graph Pad) were used for statistical significance (ns: no significative).

|  |  | SARS-CoV-2 + | Total | POS % | Mean % |
| --- | --- | --- | --- | --- | --- |
| High Risk Areas | COVID19 Hospitalization | 43 | 95 | 45,26 |  |
|  | ICU | 34 | 65 | 52,31 |  |
|  | ER | 50 | 135 | 37,04 |  |
|  | Anesthesia | 40 | 88 | 45,45 |  |
|  |  |  |  |  | 43,60 ns |
| Intermedium Risk Areas | Surgery | 79 | 175 | 45,14 |  |
|  | Oncology/Hematology | 31 | 70 | 44,29 |  |
|  | Medical areas no COVID19 | 93 | 249 | 37,35 |  |
|  | Pediatrics/Neonates | 53 | 109 | 48,62 |  |
|  | OB/GYN | 32 | 81 | 39,51 |  |
|  | Radiology | 49 | 129 | 37,98 |  |
|  | Outpatient consultation | 14 | 44 | 31,82 |  |
|  |  |  |  |  | 40,96 ns |
| Low Risk Areas | Administrative areas,<br>Clerical, Informatics,<br>Communication, Pharmacy | 37 | 67 | 55,22 |  |
|  | Laboratories | 28 | 84 | 33,33 |  |
|  | Kitchen | 18 | 47 | 38,30 |  |
|  |  |  |  |  | 41,92 ns |
| Total |  | 601 | 1438 | 41,79 |  |

**Figure 1:**
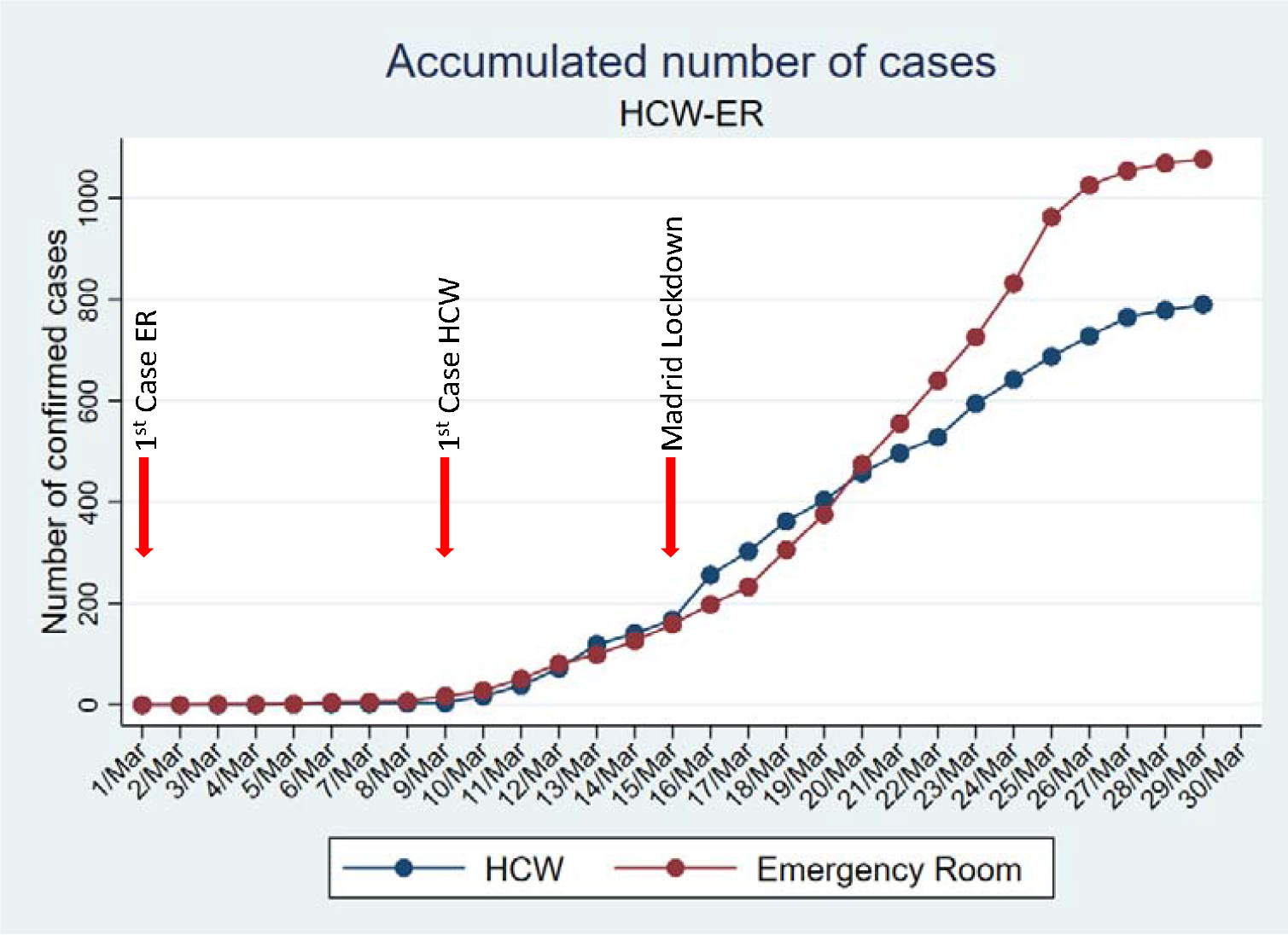
Evolution of accumulated PCR-confirmed COVID-19 cases during March 2020 at Hospital Universitario 12 de Octubre. Healthcare workers (HCW) cases evolution is compared with patients diagnosed at the Emergency Room (ER) adjusted by the same age span (20-68 years old). Highlighted by arrows the first case events and the Public Health Countermeasures adopted by the city authority.

## Discussion

The recommendation protocol from our OHSS did not include testing of asymptomatic cases, however HCW consultation was highly proactive in testing even patients with minor symptoms, therefore, a high proportion of HCW and non-sanitary personnel was tested in March 2020 during the rapid period of expansion of the epidemics in Madrid, accounting for a total of 30,6 % of the hospital employees, being this a significant representation of the institution. Most of the COVID-19 cases among the hospital HCW were mild and were managed at home under self-isolation measures, nevertheless 21 infected HCW (2,6 %) were hospitalized and 2 of them required mechanical ventilation and ICU support.

There are some differences in the rate of infection between small groups of professionals but this seems widespread throughout the whole range of risk areas. Although there could have been transmission from COVID-19 patient to HCWs, mainly at the first phase of the epidemics, there were no significant differences on the infection rates of HCW and hospital personnel that can be related to working in the groups of areas of high, intermediate and low exposure risk (Table 1). Furthermore, the evolution of cases during the same time period (March 2020) between patients attending the ER and hospital staff suggests that both groups were driven by the same dynamics (Figure 1). This experience is similar to the communicated from Wuhan and verified by the WHO Joint Mission(4) and also from recent experiences at hospitals in the Netherlands(5), where most of the infections of HCW were related to household or community contacts.

## Significance

The rate of infection of HCW, and hospital personnel, for SARS-CoV-2 during a rapid evolving epidemic wave could be an indicator of the epidemic dynamics in the community. Although we cannot exclude an additional risk factor of infection by SARS-CoV-2 due to the fact of the hospital environment, this is probably minor in the context of rapid circulation of SARS-CoV-2 in the community. This is supported by the similar proportions of positive cases among all the areas of the hospital independently of risk of exposure and also by the similar evolutive dynamics of infection as compared with the community cases attending our ER. These are clear arguments against a major factor of occupational risk and it has been also the experience of similar follow up of HCW infection in China(4) and Europe(5, 6). Exhaustive testing, such as the one carried out in our institution, covering over one third of all the workers, could be used as a reference of the proportion the population infected in the community. Since a significant proportion of COVID-19 cases can be asymptomatic and not all the hospital employees were actually tested, along with the relative lack of sensitivity of PCR testing for SARS-CoV-2 in the upper airway(7), it is highly likely that this 11,6 % is a minimum estimation of the impact of SARS-CoV-2 circulation in Madrid during the first 4 weeks of the epidemics. This is in clear contrast with the official figures circulating at national and international levels that could have grossly underestimated the actual number of cases. This has important implications to more accurately estimate the impact of the epidemics in the community and to develop health policies for containment, treatment and recovery.

## Data Availability

All data collected and analyzed for this report are available from the first author on reasonable request.

## Acknowledge

To all patients and healthcare workers involved in this pandemics and specially to those in the frontline at our institution in Madrid, Spain.

## Declaration of interest

We declare no competing interests.

All authors have seen and approved the manuscript

## Ethical approval

This study has been approved by the Institutional Internal Review

Board (IRB) of Hospital Universitario 12 de Octubre: N-CEIm: 20/169, April 5^th^ 2020.

Since all data were part of the diagnostic standard process and all information is anonymous informed consent was not considered as a requirement.

## References

1. Zhu N, et al. (2020) A Novel Coronavirus from Patients with Pneumonia in China, 2019. N. Engl. J. Med. 382(8):727–733.

2. Spiteri G, et al. (2020) First cases of coronavirus disease 2019 (COVID-19) in the WHO European Region, 24 January to 21 February 2020. Euro Surveill. 25(9).

3. https://www.thermofisher.com/es/es/home/clinical/clinical-genomics/pathogen-detection-solutions/coronavirus-2019-ncov/genetic-analysis.html

4. WHO. Report of the WHO-China Joint Mission on Coronavirus Disease 2019 (COVID-19), <https://www.who.int/docs/default-source/coronaviruse/who-china-jointmission-on-covid-19-final-report.pdf > (2020)

5. Kluytmans M, et al. SARS--CoV--2 infection in 86 healthcare workers in two Dutch hospitals in March (2020). MedRxiv preprint doi: https://doi.org/10.1101/2020.03.23.20041913

6. Hunter E, Price DA, Murphy E, van der Loeff IS, Baker KF, Lendrem D, et al. First experience of COVID-19 s creeni ng of hea lth-care workers in England. The Lancet. 2020.doi.org/10.1016/S0140-6736(20)30970-3

7. Ai T, et al. (2020) Correlation of Chest CT and RT-PCR Testing in Coronavirus Disease 2019 (COVID-19) in China: A Report of 1014 Cases. Radiology:200642.

